# Measuring weak effects in high dimensional mediation analysis

**DOI:** 10.1101/2025.01.30.25321435

**Authors:** Chunlin Li, Li Chen, James S. Pankow, Tianzhong Yang

## Abstract

Existing mediation analysis methods have often fallen short in accurately quantifying the contribution of omics mediators, particularly those with weak effects. To address this issue, we propose two new variance-based causal measures for the global mediation effect. Then, we develop a flexible and computationally efficient estimation procedure based on a mixed-effects working model. Through this approach, we are able to accurately quantify the total mediation effect and discover the weak effects that are largely mis-estimated by existing methods. The proposed approach is general and complements the existing mediation analysis methodologies by offering new perspectives on the global and weak effects.

## 1 Introduction

Many epidemiological studies have examined associations between an exposure and a health-related outcome (Mathews et al., 2015; Fernández-Sanlés et al., 2017); however, less has been done to elucidate causal pathways and biological mechanisms of the associations. Understanding these causal pathways is essential as it can enhance risk prediction, lead to the development of interventions or treatments, and guide public health policy. Mounting evidence has suggested the mediating role of molecular phenotypes, such as DNA methylation (Fujii et al., 2022) and proteins (Kim et al., 2021), measured at a massive scale, from environmental/behavioral risk factors to disease/traits. High-dimensional mediation analysis has recently become a promising tool to in investigate the underlying complex biological mechanisms.

In a mediation model, we assume

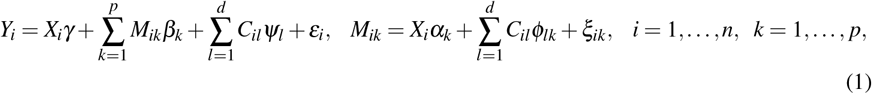

where *X*_*i*_ is an exposure variable or risk factor (e.g., age, smoking, etc.), *Y*_*i*_ is a quantitative outcome variable (e.g., blood pressure), (*M*_*i*1_, …, *M*_*ip*_) is a vector of candidate mediators (e.g., proteins, gene expression, etc.), (*C*_*i*1_, …, *C*_*id*_) is a vector of covariates (e.g., sex, race, intercept, etc.), *ξ*_*ik*_, *ε*_*i*_ are error terms, and *α*_*k*_, *β*_*k*_, *γ, ψ*_*l*_, *ϕ*_*lk*_ are unknown parameters.

### 1.1 Existing Work in High-dimensional Mediation Analysis

#### Measures of Mediation Effects

To quantify the mediation effects, most work (Zhang et al., 2016; Gao et al., 2019; Zhao and Luo, 2022) concentrates on the mean-based measures. The product and proportion measures have been widely used in the literature (MacKinnon, 2012). In (1), the product measure is defined as 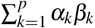. The proportion measure is characterized as the (signed) fraction of the total effect mediated by the mediators, 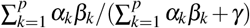, where *γ* is the direct effect, and 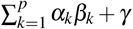 is the total effect measure. Both product and proportion measures can be viewed as mean-based measures since they rely on first-order moment relations in (1), directly summing the component-wise mediation effects. However, high-dimensional genomics mediators often exhibit effects with different directions (+/−), resulting in cancellation in a direct summation and misleading guidance toward downstream analyses.

Recognizing the weakness of mean-based measures, Huang and Pan (2016) propose to infer the quantity 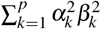. However, as a global measure of the mediation effect without direct causal interpretation, so it is mainly used as a surrogate target for hypothesis testing purposes. Yang et al. (2021) generalizes 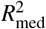 (Fairchild et al., 2009) to multivariate mediator situations. However, this measure lacks a causal interpretation. Both measures are considered variance-based measures, and this article establishes the connections between these two measures.

#### Bottom-up and Top-down Approaches

In literature, two approaches have been popular for quantifying the mediation effects: the “bottom-up” and “top-down” approaches. The bottom-up approach first identifies the active mediators and subsequently quantifies the pathway mediation effects based on the selected mediators. The bottom-up approach is able to identify the active “strong” mediators; however, in the presence of weak effects, it tends to omit these mediators, leading to underestimation of the global mediation effect. In contrast, the top-down approach estimates the aggregated mediation effects of all or a subset of mediators, which is able to capture the weak effects, offering further insights for downstream analyses. Bottom-up and top-down approaches provide complementary perspectives on mediation analysis. In this article, we focus on the top-down approach.

#### Marginal and Joint Methods

The existing methods can also be informally categorized into marginal and joint approaches. The marginal approaches focus on identifying true mediators through large-scale hypothesis testing of each mediator (Huang and Pan, 2016; Dai et al., 2022; Liu et al., 2018; He et al., 2023). Due to the large number of potential mediators and the composite nature of the hypotheses, these methods can be overly conservative. The joint approach delves into selecting mediators and estimating/testing the component-wise effects through joint regressions (Zhang et al., 2016; Guo et al., 2022; Gao et al., 2019; Zhang, 2021; Zhao and Luo, 2022; Song et al., 2020; Yang et al., 2021; Sohn et al., 2022; Perera et al., 2022). Such methods rely on sparsity as an operational assumption and may suffer from statistical inconsistency in non-sparse situations.

### 1.2 Our Contributions

This article contributes to the following aspects.

#### A New Mediation Measure

Motivated by the issue of cancellation in mean-based measures, we propose to overcome the issue by extending the *R*^2^ measure (Yang et al., 2021; Fairchild et al., 2009) to the setup of causal mediation analysis. Interestingly, this measure coincides with the one in Huang and Pan (2016) under causal and linear assumptions, thus offering a unified and new perspective on existing variance-based measures in the literature.

#### New Modeling and Estimation Strategies

We show that the proposed measure and its estimation method can capture the weak effects, thus complementing the discoveries by the mean-based measures. Our estimation procedure can accommodate different sparsity levels and different magnitudes of effects in contrast to the existing approaches. Its robustness and effectiveness are justified by a variety of simulation settings mimicking the real data scenario.

### 1.3 Organization

The rest of this article is organized as follows. In Section 2, we describe the proposed approach, including the causal *R*^2^ measure and the estimation procedure. In Section 3, we perform a comprehensive data analysis on an existing data illustrating the proposed approach. Section 4 conducts simulation studies to assess the operating characteristics of our method. Finally, Section 5 concludes the article and discusses the future directions.

## 2 Methods

In this section, we outline the proposed methods. In Section 2.1, we propose a causal *R*^2^ measure, denoted as 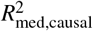, for the global mediation effect. In Section 2.2, we develop a mixed-effect working model to estimate and infer the proposed measure.

### 2.1 Measuring Global Mediation Effect

As discussed in Section 1, the existing measures of mediation effect suffer from different weaknesses. In this subsection, we adopt a causal inference framework (Pearl, 2009) and derive a mediation measure 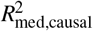 that enjoys merits desired in practice: (a) It describes the mediation effect without cancellation issue, offering a complementary view of the mediation mechanism. (b) It is intuitive with a clear causal interpretation. (c) It can be consistently estimated, regardless of sparse or dense mediators in high dimensions.

To derive the measure, we consider the model in Eq. (1). For simplicity, assume there are no mediator-mediator paths (Yuan and Qu, 2024), a common assumption in high-dimensional causal mediation analysis literature. Of note, we allow the correlation between mediators caused by some unobserved common factors.

The new measure is based on the 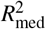 measure (Fairchild et al., 2009; Yang et al., 2021; Xu et al., 2024). The idea of 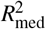 is to describe the variation of outcome *Y* explained by the exposure *X* through *M*_*k*_. Nevertheless, 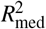 lacks causal interpretation. For example, the correlation between mediators will change 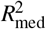, which is undesirable. To fix this issue, we would like to characterize the variability that is caused through the mediators. This is achieved by introducing stochastic interventions and defining a causal counterpart of coefficient of determination. Let us begin with a single mediator *M*_*k*_ in model (1). For (*X, M*_*k*_,*Y*), we define the causal coefficients of determination given below

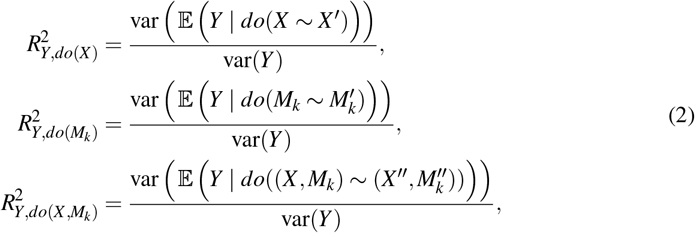

where *do*(·) is the do-operator in causal inference (Pearl, 2009), and 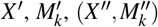 are independent copies of *X*, *M*_*k*_, (*X, M*_*k*_), respectively. The causal coefficient of determination is closely connected to the causal entropy (Simoes et al., 2023). As such, for *M*_*k*_ we define the causal counterpart of 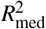 (Fairchild et al., 2009) as 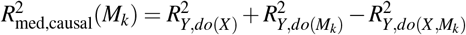. For *M*_1_, …, *M*_*p*_, when no mediator-mediator path exists, the measure for the total mediation effect is given by 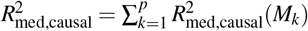. Assume the following identifiability conditions:

1. No unmeasured confounding of the exposure-mediator effect.
2. No unmeasured confounding of the mediator-outcome effect.
3. No unmeasured confounding of the exposure-outcome effect.
4. No mediator-outcome confounder that is itself affected by the exposure.

We have the following result.

#### Theorem 1.

*In Eq*. (1), *if the identifiability conditions are satisfied, then we have*

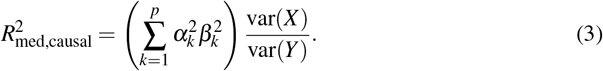

The new measure 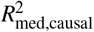 unifies two existing measures, overcoming their weaknesses: it extends 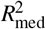 under the causal framework and provides 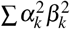 an accessible interpretation. Thus, our derivation offers valuable insight for choosing a proper mediation measure in practice. Note that causal *R*^2^ can be larger than one, although, in practical scenarios, this value is usually between zero and one, as the signal-to-noise ratio in genomics studies is typically quite low.

In addition to 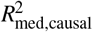, we also define 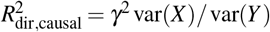, and a relative measure

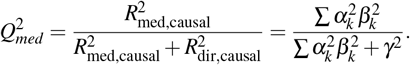

This measure quantifies the proportion of the mediation effect among the total effect and is range from 0 to 1.

### 2.2 Mixed Effect Working Model

This subsection develops a working model for estimating 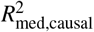 as well as 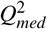.

In high-dimensional situations, to identify true mediators, existing methods mainly focus on settings where the signals are strong but sparse. For weak signals 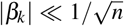, existing methods yield either diminishing power or inconsistent estimation. Despite being too weak to be accurately estimated individually, non-sparse faint signals can contribute significantly to global mediation effects. To accommodate different sparsity levels and signal strengths, we model the sparse strong signals as fixed effects and the non-sparse weak signals as random effects.

To describe the strategy, we partition the set of candidate mediators into strong and weak mediators. Let 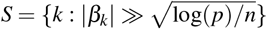 be the set of strong mediators. We assume {*α*_*k*_} is fixed. For *k* ∈ *S*, we assume *β*_*k*_ are fixed. For *k* ∈ *S*^*c*^, assume 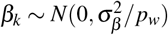 independently, where *p*_*w*_ = *p* −|*S*|. Thus, by the law of large numbers, when the number of mediators *p* ⪢ *n* is large, we have

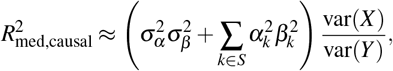

where 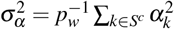 and 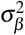 is the variance component of random effects. On this basis, we fit a high-dimensional linear mixed effect model to estimate 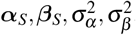. Specifically, we (a) identify the sparse fixed effects ***α***_*S*_, ***β***_*S*_ based on the de-correlation techniques (Wang et al., 2016; He et al., 2024), and (b) fit the variance components 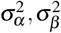 of random effects using a modified procedure based on the average information restricted maximum likelihood (AI-REML). We summarize the proposed procedure as follows:

- **Step 1. (***K***-fold data splitting)** The original sample 𝒟 is randomly split into *K* equal subsamples 𝒟^(1)^, …, 𝒟^(*K*)^.
- **Step 2. (Cross fitting)** Let 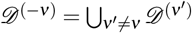.
  - *Step 2.1*. In 𝒟^(−*ν*)^, perform screening on *α* with FDR control, denoted as 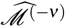. Using 𝒟^(*ν*)^, estimate 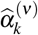 and 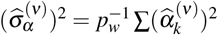, where 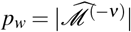.
  - *Step 2.2*. In 𝒟^(*ν*)^, identify strong mediators 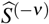 using decorrelated methods (Wang et al., 2016; He et al., 2024) or other methods like HDMT (Dai et al., 2022).
  - *Step 2.3*. Using 𝒟^(*ν*)^, fit mixed effect model using ML for 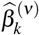 and REML for 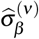
- **Step 3. (Final estimate)** Compute

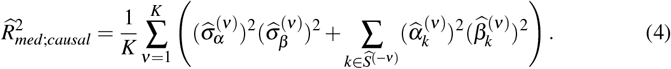

Importantly, it overcomes the issue of error accumulation in existing work by estimating 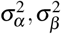 instead of individual component-wise (non-sparse) effects.

## 3 Application to the Multi-Ethnic Study of Atherosclerosis (MESA)

MESA is a longitudinal cohort study designed to investigate risk factors for cardiovascular disease (CVD) and the progression from subclinical to clinical CVD. MESA researchers study a diverse, population-based sample of 6,814 asymptomatic men and women aged 45–84. As part of the NHLBI Trans-Omics for Precision Medicine (TOPMed) Whole Genome Sequencing Program, omics data of a subset of MESA participants (921 subjects) were collected and made available through dbGap (phs001416.v3.p1). We applied our algorithm to explore the role of 1318 proteins measured from venous blood at exam 1 in the pathway from age to systolic blood pressure (SBP). The proteins were all assayed by the Broad Institute using a SOMAscan HTS Assay 1.3K for plasma. We extracted race, sex, age, height, weight, and smoking status (ever vs. never) at exam 1, and SBP from exam 2. Due to the missingness in the data (*<* 2% missing for covariates in exam 1, ~ 5% missing for SBP in exam 2), we used the MICE algorithm in R for multiple imputation. Additionally, we inverse norm transformed the protein levels, from which the first thirteen PCs were extracted to reduce residual confounding (Xu et al., 2024). We used virtual examination of the observed correlation matrix among proteins after adjusting for these PCs (Fig. 1) and the number of PCs was decided based on the scree plot. We examined whether plasma proteins measured at exam 1 may mediate the association between age and exam 1 and SBP at exam 2, adjusting for weight, height, PCs, race, and sex. In the pathway from age to SBP, we started by exploring *α*_*k*_ and *β*_*k*_, where we observed strong evidence of the existence of weak bidirectional (+/−) effects (Fig. 2).

**Figure 1:**
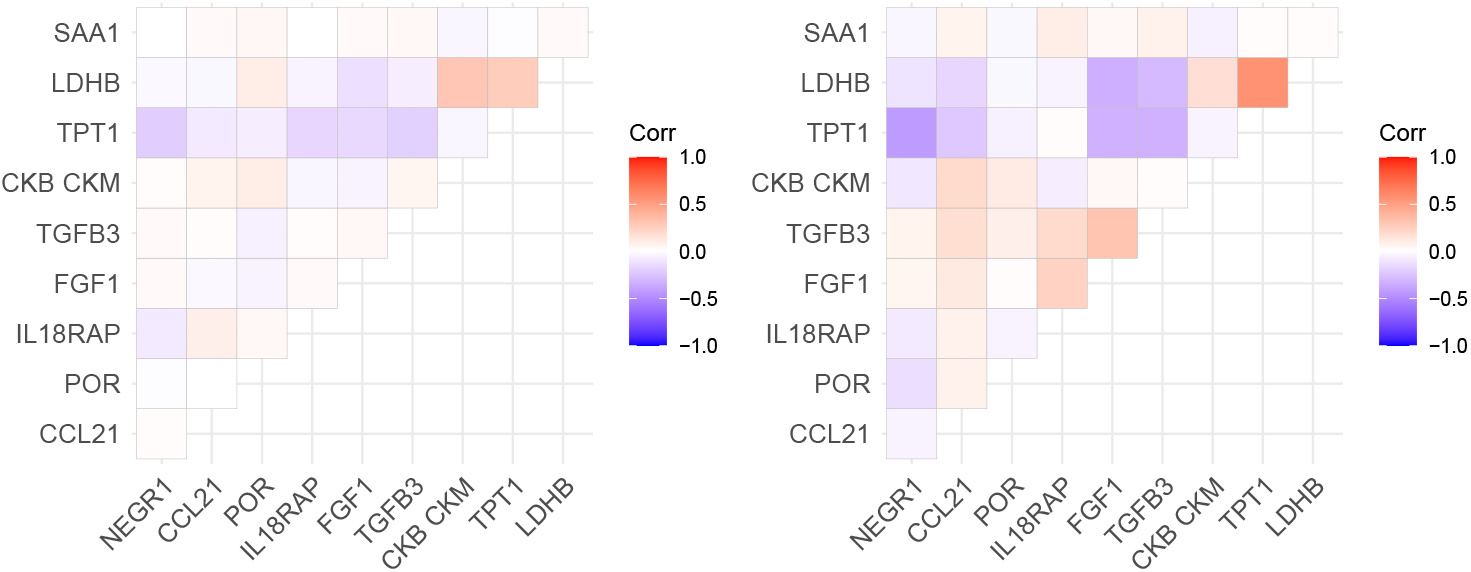
Correlation among 10 randomly chosen proteins before (Right) and after adjusting for PCs (Left).

**Figure 2:**
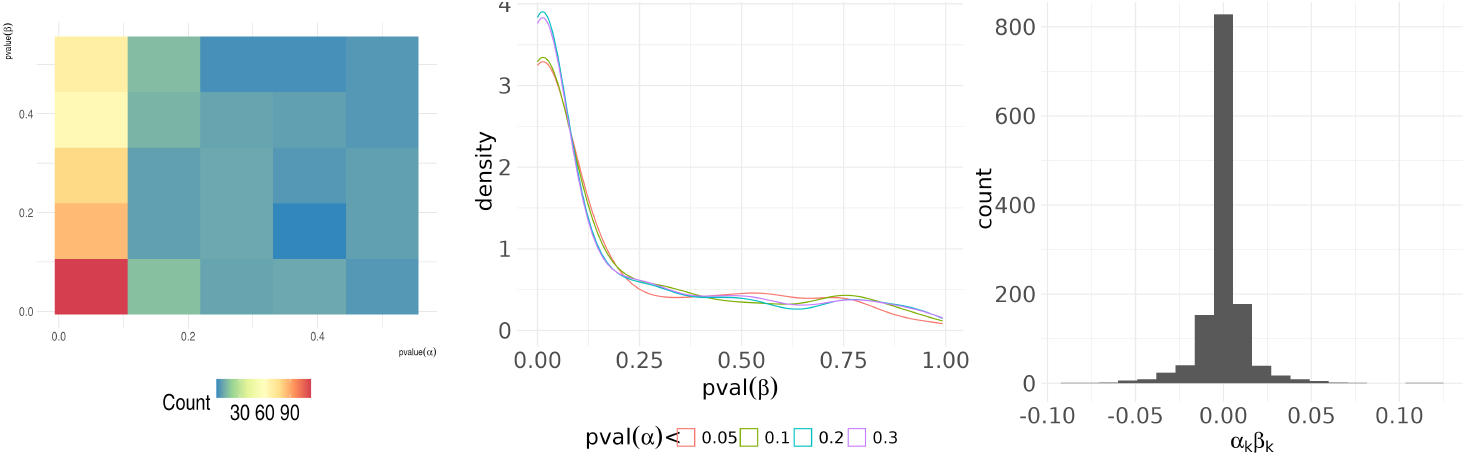
The *α*_*k*_ and *β*_*k*_ estimates for the path age-protein-SBP in MESA. Left: Heatmap of pairwise p-values of *α*_*k*_ and *β*_*k*_; Middle: Density plot of p-values of *β* with p-values of *α* less than 0.05, 0.1, 0.2, and 0.3; Right: Histogram of *α*_*k*_*β*_*k*_ of all proteins.

We estimated the 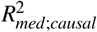 and 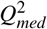 for all of the 1317 proteins, 91 proteins belong to the response to stress pathway (GO:0006950), and 123 proteins belong to the circulatory system process (GO:0003013). Acute and chronic stress has been well recognized to be closely related to blood pressure regulation through various mechanisms (Gasperin et al., 2009), whereas the circulatory system process is a broad category involving blood circulation, heart process, vascular process, etc. Genes associated with blood pressure identified in a genome-wide association study were found to be enriched for this pathway (Zhang et al., 2015). Note that there was an overlap of 18 genes between the response to stress pathway and the circulatory system process.

Following the proposed estimation step, we used HDMT to identify strong signals and report results in Tables 1 - 2. REN, the renin protein that generates angiotensin I from angiotensinogen in plasma, was identified in the effects from and age to SBP. REN was included in the circulatory system process but not in the response to stress pathway. In addition to the selected proteins, we discovered a range of ratio of variance of outcome due to weak mediators to strong mediators from age to SBP (WS ratio in Table 1). Compared to HIMA (Zhang et al., 2016), HDMA (Gao et al., 2019), our 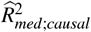 captured much higher variance mediated (Table 2). Together, these observations suggested a large amount of mediation effect was missed by existing methods. In addition, the circulartory system process pathway showed higher contribution to the mediation effect than the response to stress pathway (Table 1). We also observed very different results of the product measure for total mediation effect across these competing methods, i.e., HDMA, HIMA, and HILMA (Zhou et al., 2020).

**Table 1:** Empirical results from age to SBP in MESA. WS ratio is the ratio of variance of outcome due to weak mediation effects and strong mediation effects, where the proteins with strong effects were selected by the HDMT method.

| # proteins | pathway | $R^2_{med;causal}$ | $Q^2_{med}$ | WS ratio | Selected protein |
| --- | --- | --- | --- | --- | --- |
| 1317 | all proteins | 0.0061 | 0.89 | 10.95 | REN |
| 91 | Response to stress | 0.0010 | 0.52 | ∞ | |
| 123 | Circulatory system process | 0.0015 | 0.64 | 4.82 | REN,STC1 |

**Table 2:**
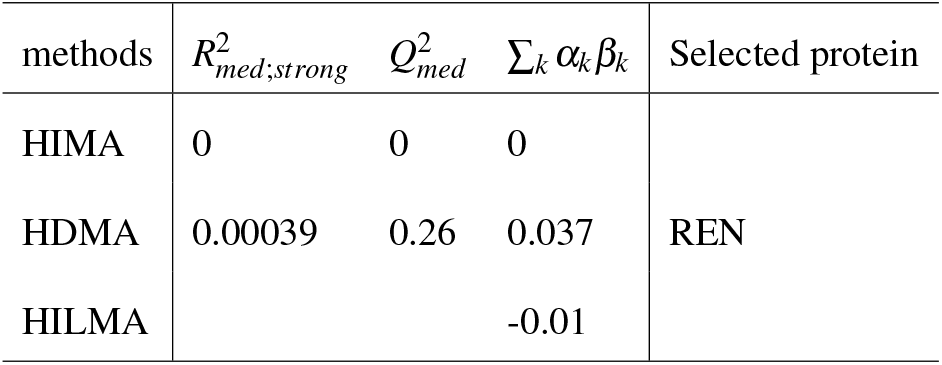
Comparative methods for all proteins from Age to SBP. Bonferroni correction was applied to the p-values of selected proteins for HIMA and HDMA.

## 4 Simulation Studies

This section examines the proposed analytical procedure through simulations.

### Evaluating 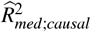 with different effect strength

We evaluated the bias and standard deviation of 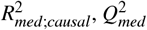 under three settings of different combinations of strong and weak effects: (a) strong and sparse mediation effects only, (b) weak and non-sparse effects only, and (c) both strong sparse and weak and non-sparse effects. For each scenario, we estimated 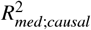 taking into the strong effects only, weak effects only, and both, denoted as 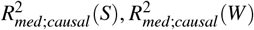, and 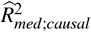, respectively. We also compared with 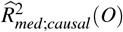 as if 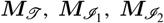 and 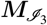 are known. The simulation data generating equations followed Eq. (1). We set *p* = 2000, *n* = 1500, *γ* = 0.4, *ε* ~ *N*(0, 0.5^2^) and *X* ~ *N*(0, 1) across the three scenarios. We let *U* ~ *N*(0, *I*_*N*×2_), ***α***_0_ ~ *N*(0, 0.3) and ***ξ***_0_ ~ *N*(0, *I*_*N*×1_). Each scenario was replicated 200 times.

- **Scenario 1** with strong and sparse signal: There were 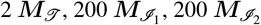 and the rest were 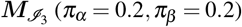. True mediators had strong effects: *α* = [1.5, 1.5], *β* = [0.2, −0.2]; Non-mediators with weaker effects: 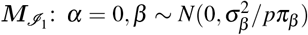 where 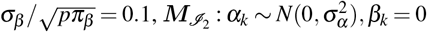, where *σ*_*α*_ = 0.7, and 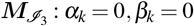;
- **Scenario 2** with weak and non-sparse signal: There were 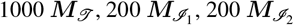, and the rest were 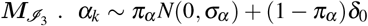 and 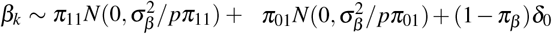, where 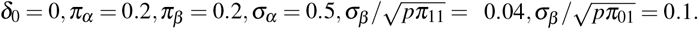.
- **Scenario 3** with both: There were 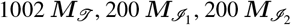, and the rest were 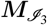. Five of ***M***_𝒯_ are of strong effects: *α* = [1.5, 1.5], *β* = [1.5, −1.5], where the rest were of weak effects following the same distribution as in **Scenario 2**.

Note that we assume weaker effects on ***β*** than ***α*** based on observation of the relative scale in real data. When estimating 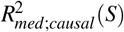 and 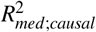, we used the HDMT method (Dai et al., 2022) to identify strong signals because it does not assume sparsity. When estimating 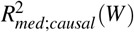, we ignored the potential strong signals and directly fit a mixed effect model.

Table 3 summarizes the result. Our proposed estimator 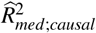 exhibited a bias level comparable to that of the oracle estimator 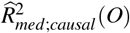 across all three settings, although it had larger standard error. As expected, 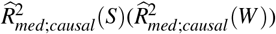 only had low bias when there were only strong(weak) signals. *Q*^2^ was annotated and calculated similarly.

**Table 3:**
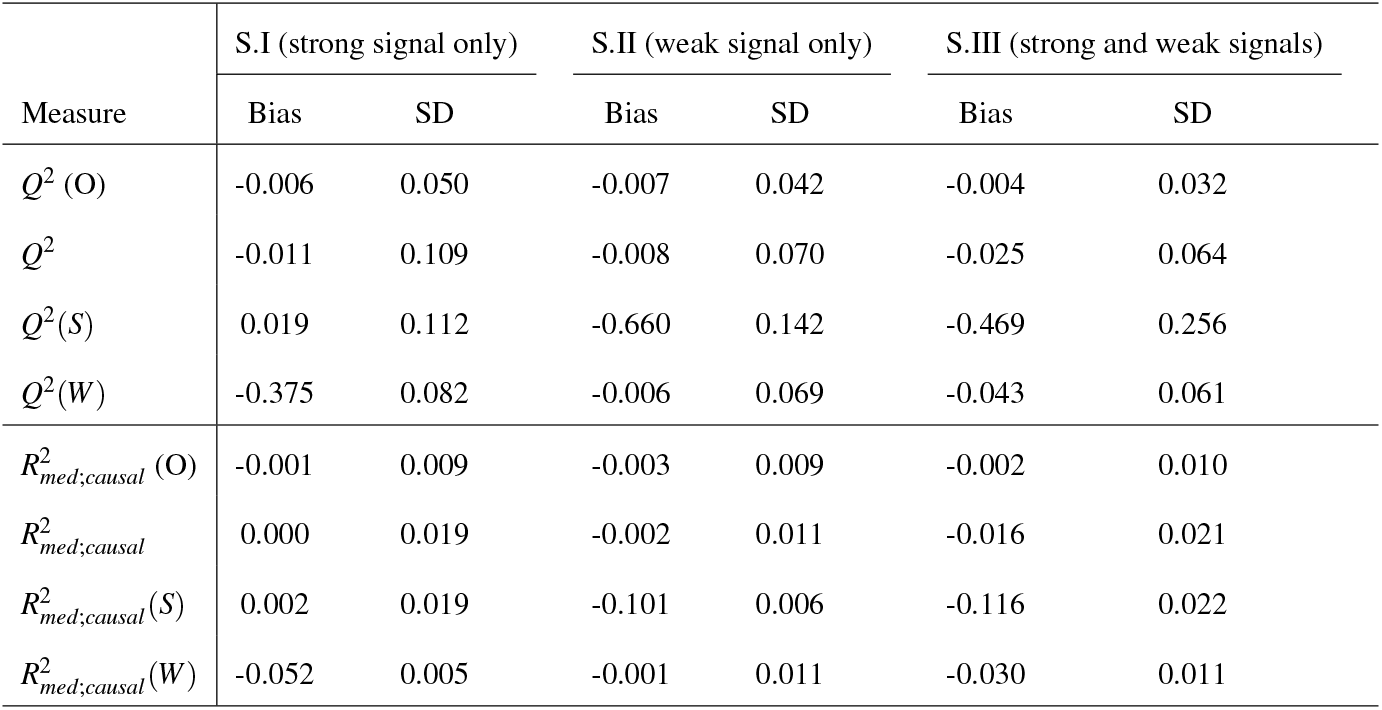
Simulation results to evaluate mediation with different stengths. **SD** refers to the empirical standard deviation of bias. True 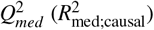 in Scenarios 1, 2, 3 are 0.5294 (0.06218), 0.7715(0.1055) and 0.8182 (0.1395), respectively.

### Evaluating consistency of 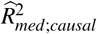

In **Scenario 2**, we further evaluated the performance of 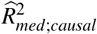 varying *n* at 500, 1000, and 1500. We fixed the total number of putative mediators at 2000 and keep the same 1000 true mediators ***M***_𝒯_. We varied the number of 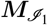 and 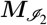 at S1 (500, 200), S2 (800,200), S3 (200,500) and S4 (200, 800), respectively.

Figure 3 demonstrates that both bias and variance decrease as the sample size increases, suggesting the consistency of the proposed estimator.

**Figure 3:**
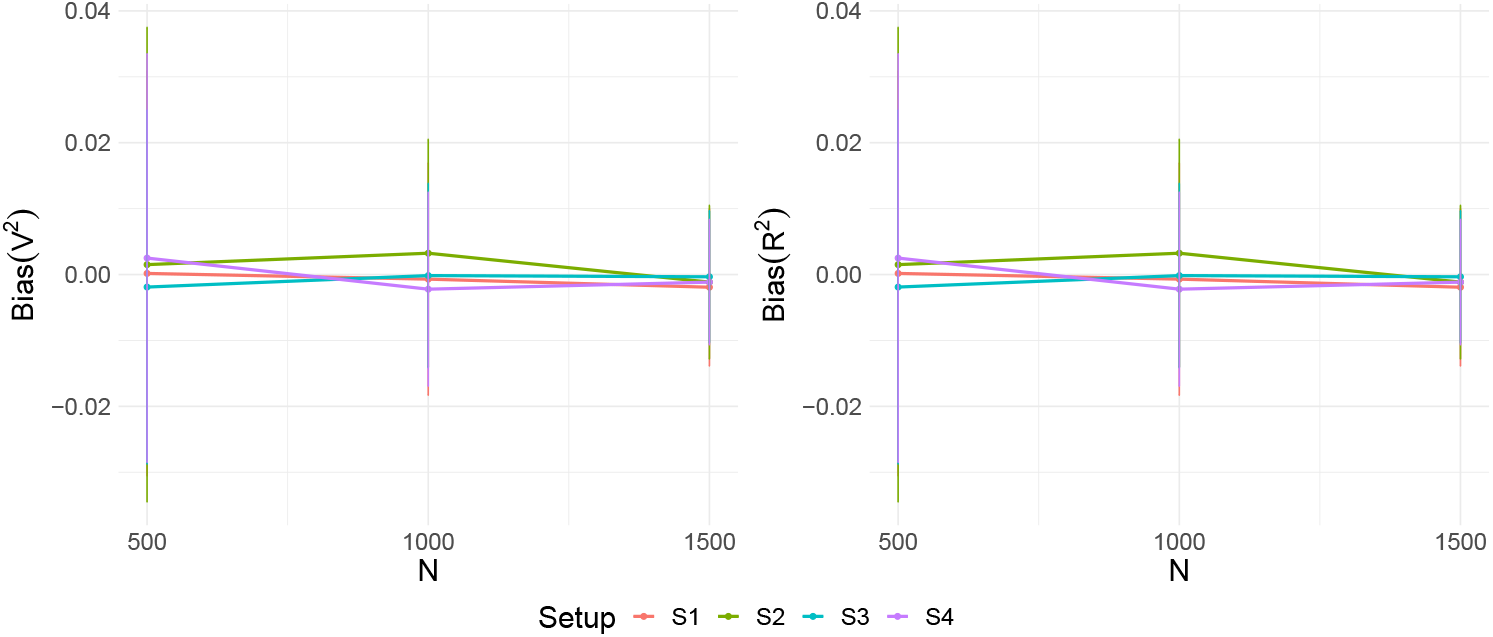
Evaluation of consistency of *Q*^2^ (Left) and 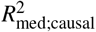 (Right) under different types and numbers of non-mediators

## 5 Discussion

This article proposes a new *R*^2^ measure, denoted as 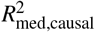, together with a relative measure 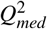. In the linear mediation model, under standard causal assumptions, 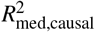 coincides with the existing measure. Thus, it provides a unified interpretation of existing variance-based measures. Built upon this, we develop a mixed-effect working model, separating strong and weak effects, where strong effects are modeled as fixed effects while weak effects as random effects. This model is suitable for capturing the weak signals when they collectively contribute a significant portion to the outcome. Through the application to the MESA data, we find that weak effects are common in real data while existing methods largely ignore them. Our approach thus provides a complementary measure for high-dimensional mediation analysis.

Our approach is general and applicable to many other studies. We plan to extend this measure and estimation method to binary outcomes, including the case-control designs.

## Data Availability

No new data is produced.

https://www.ncbi.nlm.nih.gov/projects/gap/cgi-bin/study.cgi?study_id=phs001416.v3.p1

## 6 Acknowledgement

MESA and the MESA SHARe project are conducted and supported by the National Heart, Lung, and Blood Institute (NHLBI) in collaboration with MESA investigators. Support for MESA is provided by contracts HHSN268201500003I, N01-HC-95159, N01-HC-95160, N01-HC-95161, N01-HC-95162, N01-HC-95163, N01-HC-95164, N01-HC-95165, N01-HC-95166, N01-HC-95167, N01-HC-95168, N01-HC-95169, UL1-TR-001079, UL1-TR-000040, UL1-TR-001420, UL1-TR-001881, and DK063491. Dr. Pankow’s effort in MESA was supported by contract 75N92020D00006 from the National Heart, Lung, and Blood Institute. Dr. Yang was supported by St. Baldrick Career Award and NIH grant R01AG074858. The authors acknowledge the Minnesota Supercomputing Institute (MSI) at the University of Minnesota for providing resources that contributed to the research results reported within this paper.

